# Understanding dynamics of respiration amongst sexes: who breathe more efficiently?

**DOI:** 10.1101/2023.03.28.23287803

**Authors:** Dev Himanshubhai Desai, Prahasth Dave, Anita Verma, Neeraj Mahajan

## Abstract

**Introduction:** Respiration is a complex phenomenon requiring diaphragm, inter-costal muscles and other supporting structures. Contemplating the anatomical & physiological differences between males and females, it is essential to know how the respiratory system works in both of them. No such other study has been conducted in an Indian setup, which guided us to take up this topic.

**Methodology:** Total of N= 216 (Males 63, Females- 153) student were enrolled. All the participants were between the age of 17-19. Their data of Tidal volume, inspiratory reserve volume, expiratory reserve volume, maximum expiratory pressure and their vital capacity both sitting and standing were gathered and analysee.

**Results:** Tidal volume was in males 553±56 ml and 666±60 in females(p-value = 0.031). IRV was in males 2103±139 ml and 1717±99 in females(p-value>0.0001). ERV was in males 1638±113 ml and 1323±65 in females (p-value>0.0001). VC Standing was in males 3947±155 ml and 3278±105 in females(p-value>0.0001). VC sitting was in males 3492±151 ml and 2743±107 in females(pvalue>0.0001). MEP was in males 90±8 mmHg and 64±6 mmHg in females(p-value>0.0001).

Range of Pearson correlation coefficient for all=(+0.2)-(−0.2).

**Conclusion:** Tidal volume was found to be higher in females than in males. Vital capacity was higher in males than in females by 700 ml in both position and vital capacity was higher by 500ml in standing than in sitting in both males and females. Body mass index weakly correlatable positively or negatively with all parameters. MEP was found to be higher in males but was weakly correlated negatively with BMI.

## Introduction

Respiration, a lifelong life-sustaining process. Respiratory process started from the single cellular organisms that first came on the earth and has evolved with the evolution of the life itself. From, mere taking of oxygen in Aerobic Bacteria (1) to having few molecules of hemoglobin to hold oxygen in lower single cellular organism, and from using skin to respire in annelids (2) to using gills by fishes (3), and lungs in mammals (4) evolution has increased the respiratory power tremendously.

Humans breathe their whole life, even while sleeping or when sick. In different conditions, the respiration process changes and that can actually change the outcome of the event (5) from death to life or life to death. Respiratory power is actually a quantifiable and countable entity with the help of different parameters namely Tidal Volume (6) Inspiratory Reserve volume (7), Expiratory Reserve volume (8), Vital Capacity (9) & Maximum Expiratory pressure (10). This all parameters are measured by a technique called Spirometry (11) and the apparatus used for this is called Spirometers (12)

Cardiac events have a significant impact on respiration. A person’s weight and BMI also have a significant impact on the parameters. It has been found extensively that obesity and adipose fat collection reduces the capacities of lungs to hold air and respire. (13)

A lot of studies have been conducted in many countries trying to normalize the average range of values of the volume and capacities of lungs but the multifactorial aspect affecting the lung has caused a drastic difference between these. Here, we are trying to normalize the values of lung capacity in young adolescents from a very diverse population group to increase our precision, so we can be as close as to the true mean.

### Methodology

A cross sectional study was designed to collect data. Students studying at Smt. NHL Municipal Medical College were enrolled as study participants. Physiology Department of the college collects data of the students as their normal practical curriculum. The data was acquired from the department with the permission of head of the department

All the data gathered was digitalized and was used to analyze the outcomes.

### Study design

Cross Sectional record based Study

### Study population

after excluding students by the exclusion criteria, total of 216 students were enrolled from which 63 were males and 153 were female student. All of them were of age group of 17-19

Inclusion criteria was students who are willing to give consent at the time of the data recording and who did not suffer from any respiratory diseases. While students not willing to consent or with respiratory disease were excluded. Also students feeling sick or unwell were excluded.

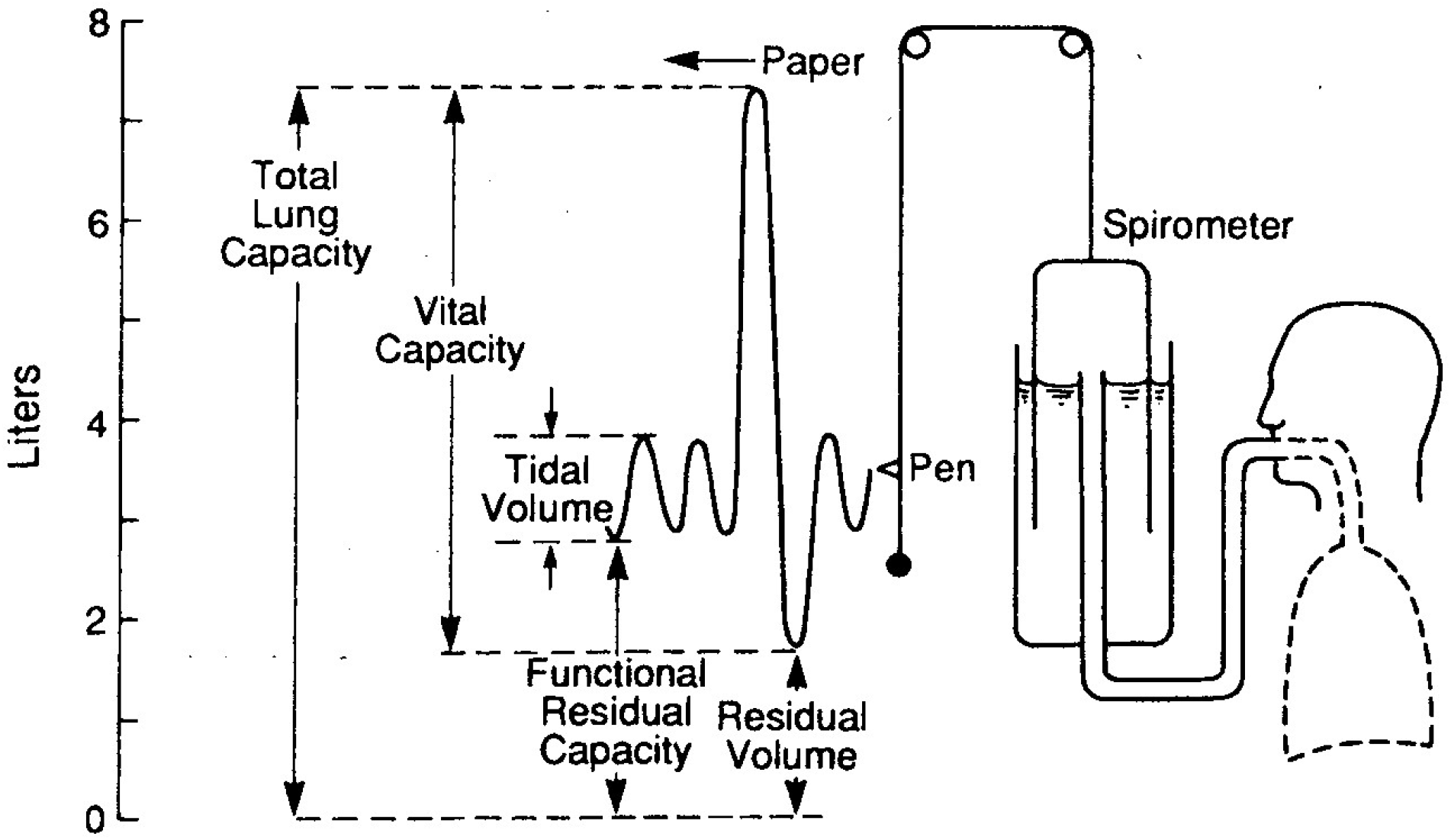

The data the students was recorded for the following parameters:-Tidal Volume (TV), Inspiratory reserve volume (IRV), Expiratory reserve volume (ERV), Vital Capacity (VC), Sitting VC, Standing VC, Maximum expiratory Pressure (MEP)

Statistical tests of Average, Standard deviation, Standard error of mean, Confidence interval (CI_95_) analysis done, Pearson correlation (R value), Statistical significant test (unpaired t test) (p value) with Charts and tables and Comparative analysis across ranges were done using Microsoft excel and SPSS20.

## Results

1. Tidal Volume (TV) (Figure 1, Table 1)

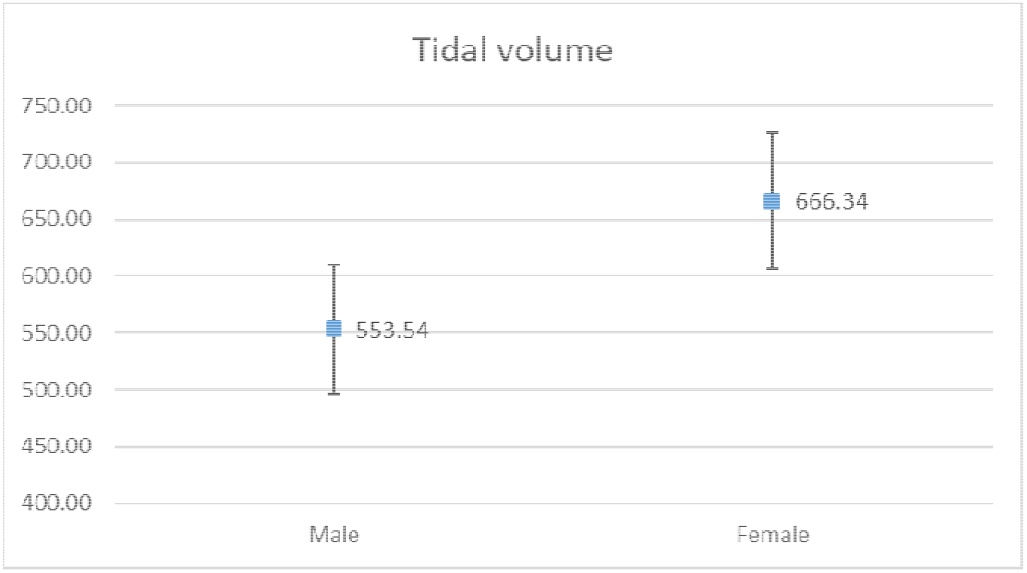

2. Inspiratory Reserve Volume (IRV) (Figure 2, Table 2)

1. Tidal Volume (TV) (Figure 1, Table 1)
|  | Male | Female |
| --- | --- | --- |
| Average | 553.54 | 666.34 |
| Standard Deviation | 228.86 | 378.35 |
| N | 63.00 | 153.00 |
| SEM | 28.83 | 30.59 |
| CI95 | 56.51 | 59.95 |
| P value | 0.031 |  |

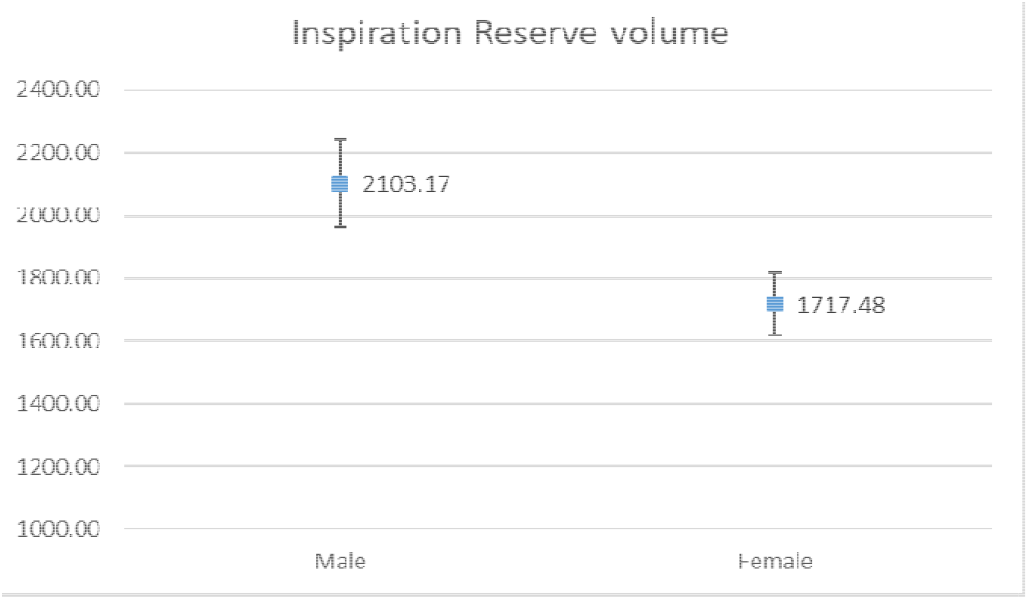

3. Expiratory Reserve Volume (IRV) (Figure 3, Table 3)

2. Inspiratory Reserve Volume (IRV) (Figure 2, Table 2)
|  | Male | Female |
| --- | --- | --- |
| Average | 2103.17 | 1717.48 |
| Standard Deviation | 565.89 | 626.98 |
| N | 63.00 | 153.00 |
| SEM | 71.30 | 50.69 |
| CI95 | 139.74 | 99.35 |
| P value | <0.0001 |  |

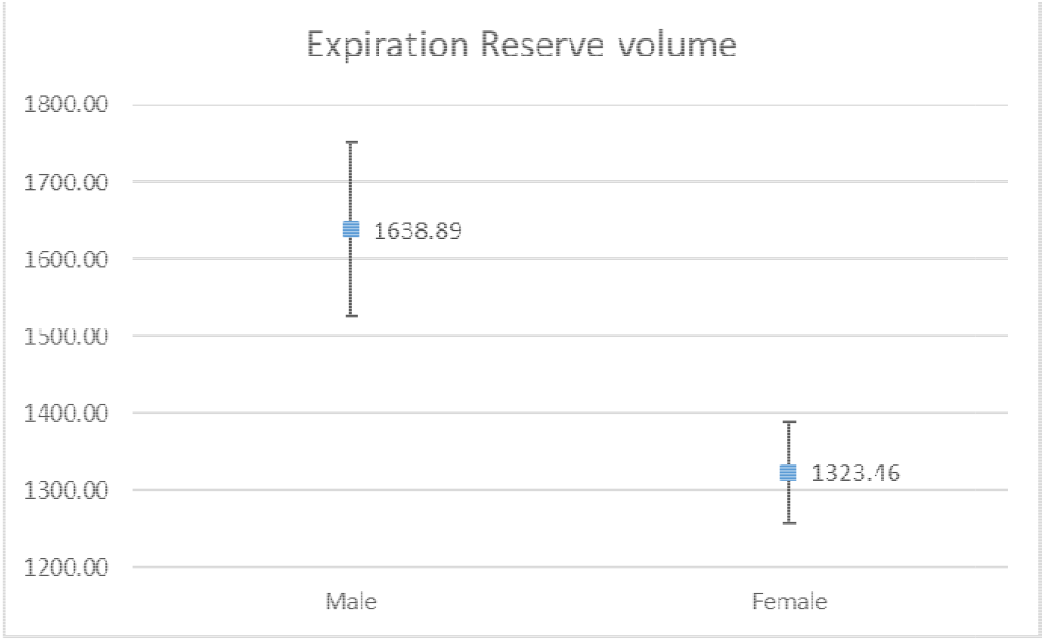

4. Vital Capacity Standing (VC standing) (Figure 4, Table 4)

3. Expiratory Reserve Volume (IRV) (Figure 3, Table 3)
|  | Male | Female |
| --- | --- | --- |
| Average | 1638.89 | 1323.46 |
| Standard Deviation | 459.53 | 414.99 |
| N | 63.00 | 153.00 |
| SEM | 57.89 | 33.55 |
| CI95 | 113.47 | 65.76 |
| P value | <0.0001 |  |

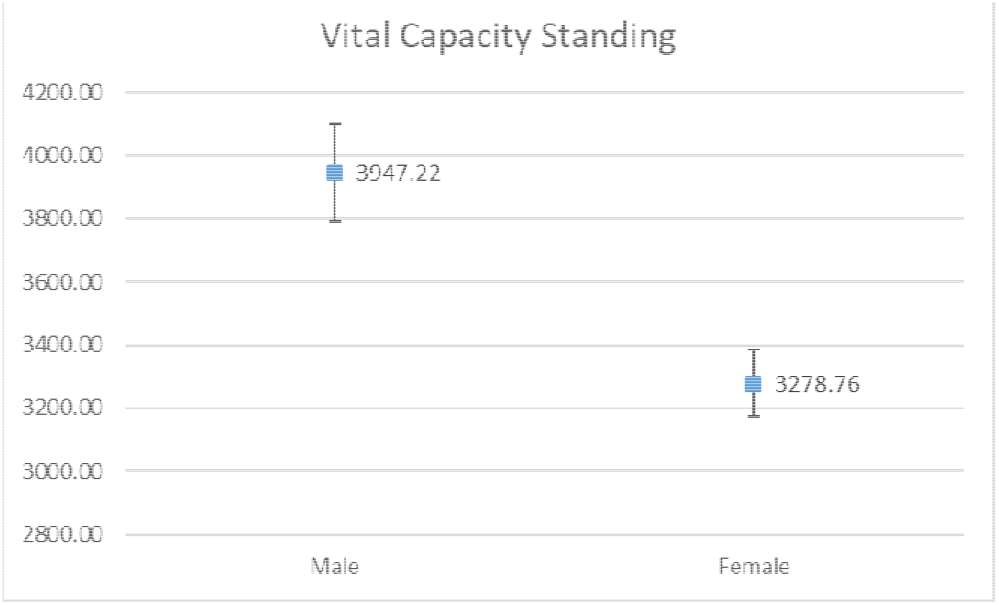

5. Vital Capacity Sitting (VC sitting) (Figure 5, Table 5)

4. Vital Capacity Standing (VC standing) (Figure 4, Table 4)
|  | Male | Female |
| --- | --- | --- |
| Average | 3947.22 | 3278.76 |
| Standard Deviation | 631.69 | 663.84 |
| N | 63.00 | 153.00 |
| SEM | 79.59 | 53.67 |
| CI95 | 155.99 | 105.19 |
| P value | <0.0001 |  |

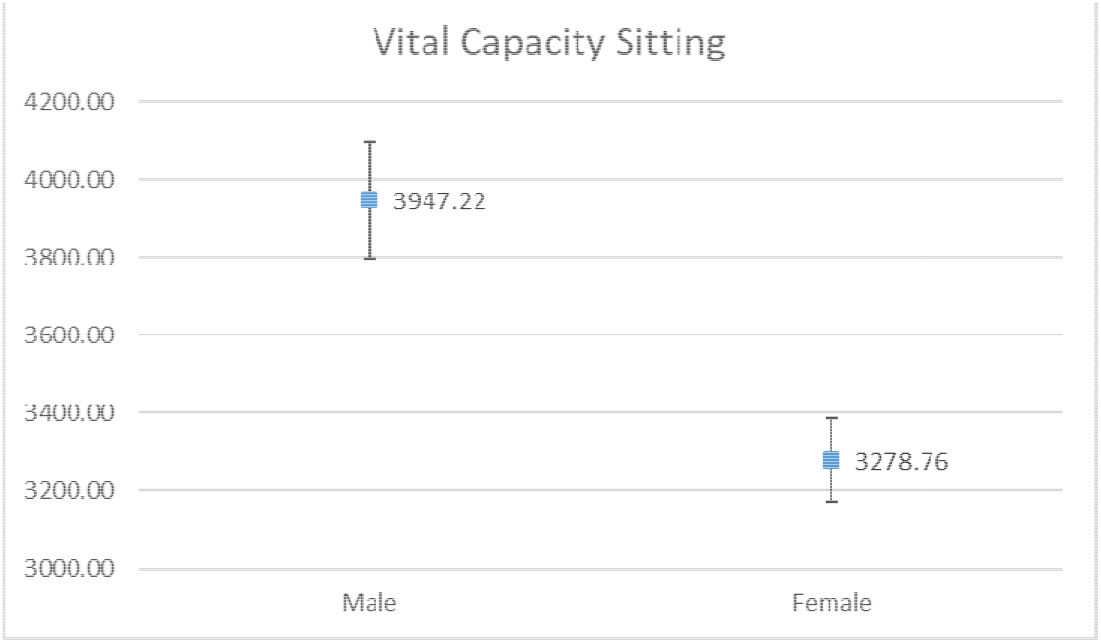

6. Maximum Expiratory Pressure (MEP) (Figure 6, Table 6)

5. Vital Capacity Sitting (VC sitting) (Figure 5, Table 5)
|  | Male | Female |
| --- | --- | --- |
| Average | 3492.86 | 2743.03 |
| Standard Deviation | 612.49 | 676.42 |
| N | 63.00 | 153.00 |
| SEM | 77.17 | 54.69 |
| CI95 | 151.25 | 107.18 |
| P value | <0.0001 |  |

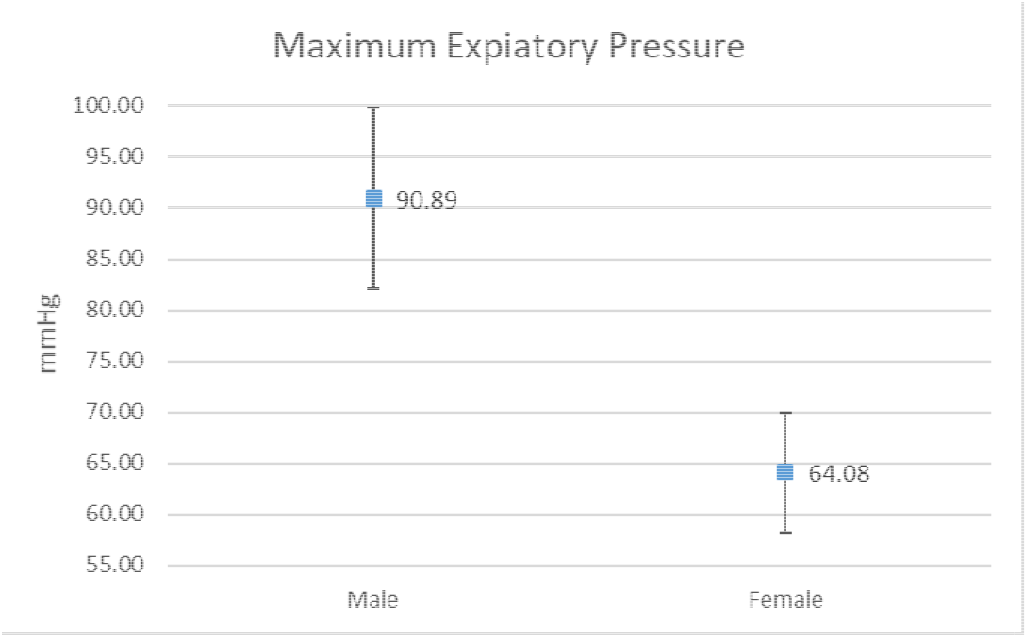

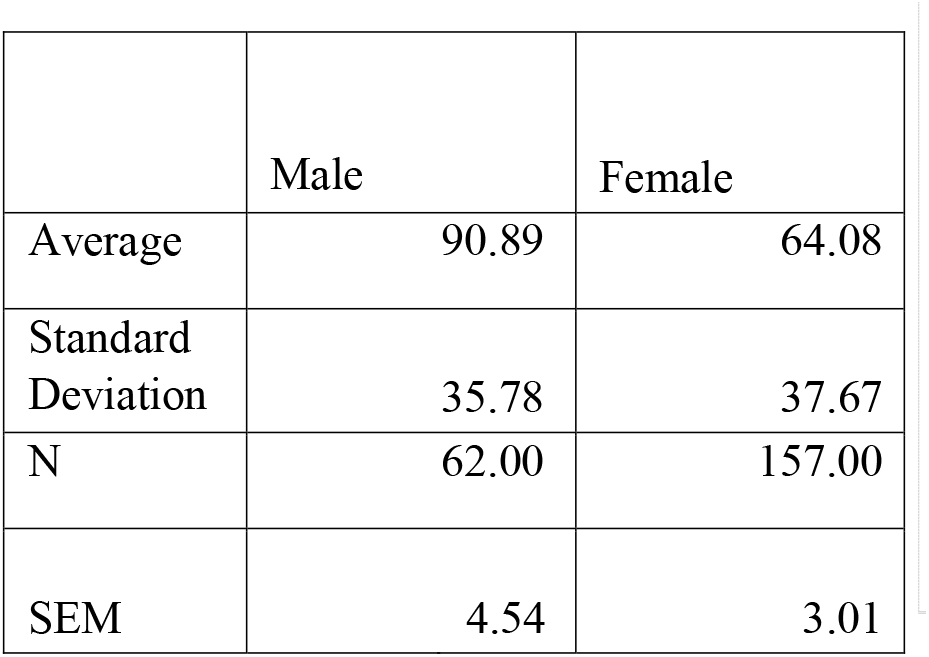
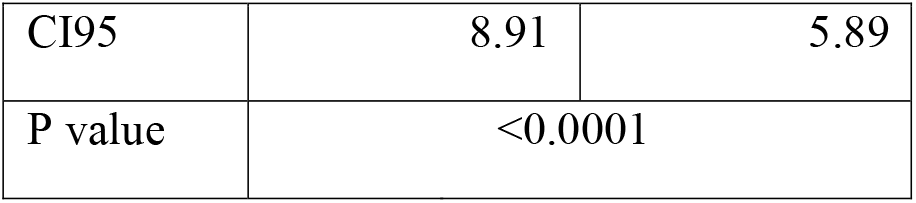
7. Comparison of Pearson Correlations of BMI and all respiratory parameters between males and females
  a. Tidal volume (Table 7, Figure 8, Figure 9)

6. Maximum Expiratory Pressure (MEP) (Figure 6, Table 6)
|  | Male | Female |
| --- | --- | --- |
| Average | 90.89 | 64.08 |
| Standard Deviation | 35.78 | 37.67 |
| N | 62.00 | 157.00 |
| SEM | 4.54 | 3.01 |
| CI95 | 8.91 | 5.89 |
| P value | <0.0001 |  |

7. Comparison of Pearson Correlations of BMI and all respiratory parameters between males and females a. Tidal volume (Table 7, Figure 8, Figure 9)
| R- value ( vs BMI) | Males | Females |
| --- | --- | --- |
| TV | -0.0587 | -0.2350 |

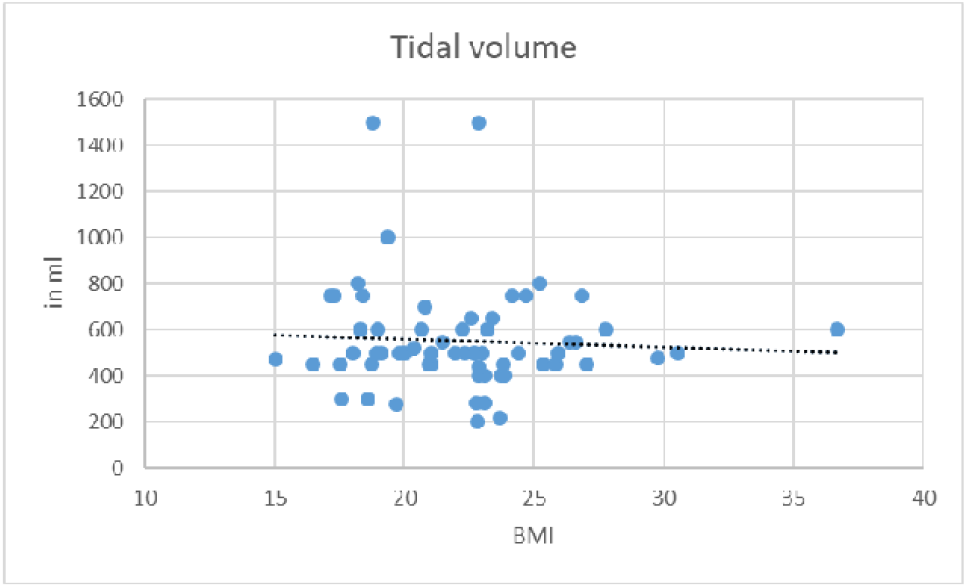

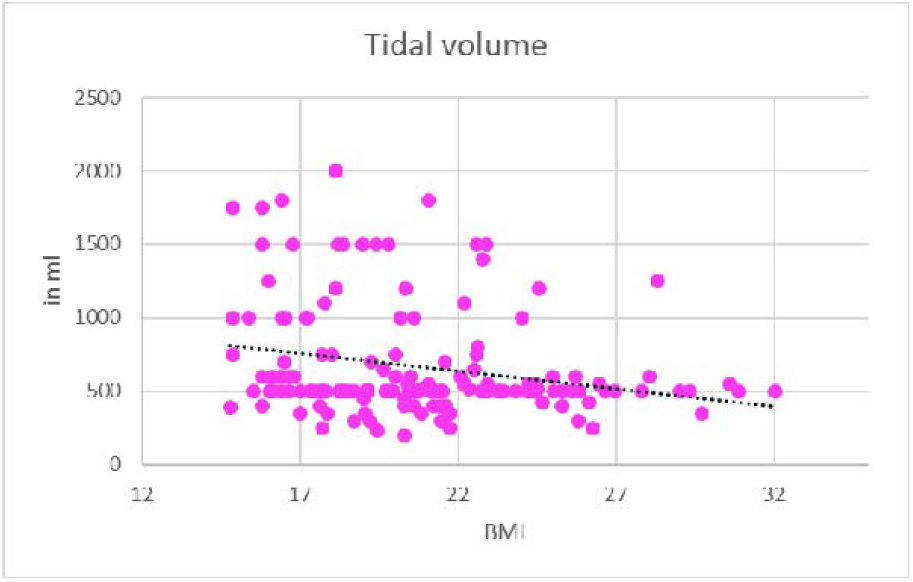
  b. Inspiratory reserve volume (Table 8, Figure 10, Figure 11)

| R- value ( vs BMI) | Males | Females |
| --- | --- | --- |
| IRV | 0.1819 | 0.1492 |

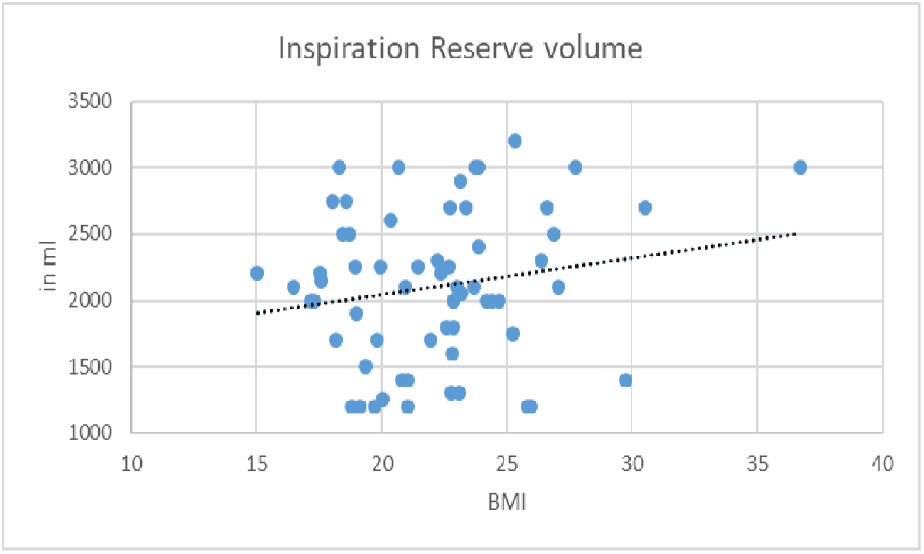

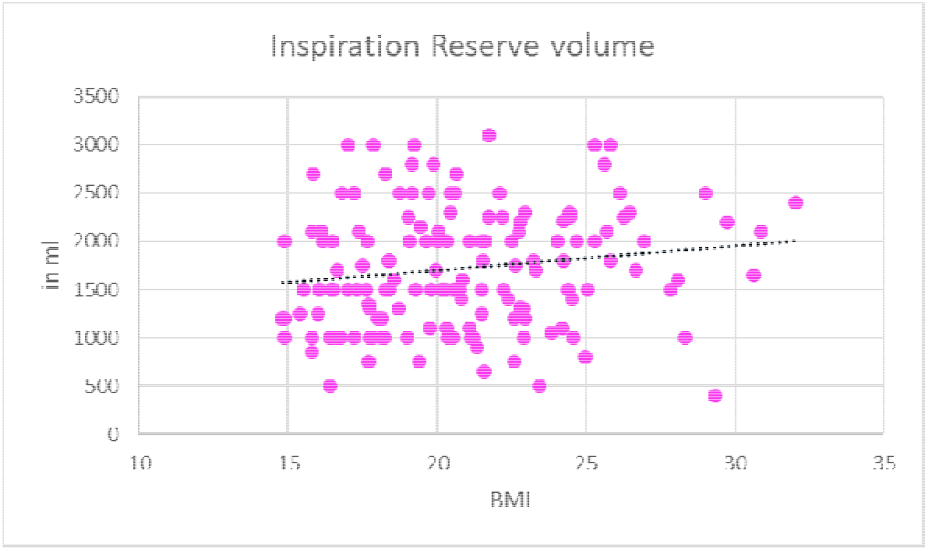
  c. Expiratory Reserve Volume (Table 9, Figure 12, Figure 13)

| R- value ( vs BMI) | Males | Females |
| --- | --- | --- |
| ERV | 0.1277 | -0.0465 |

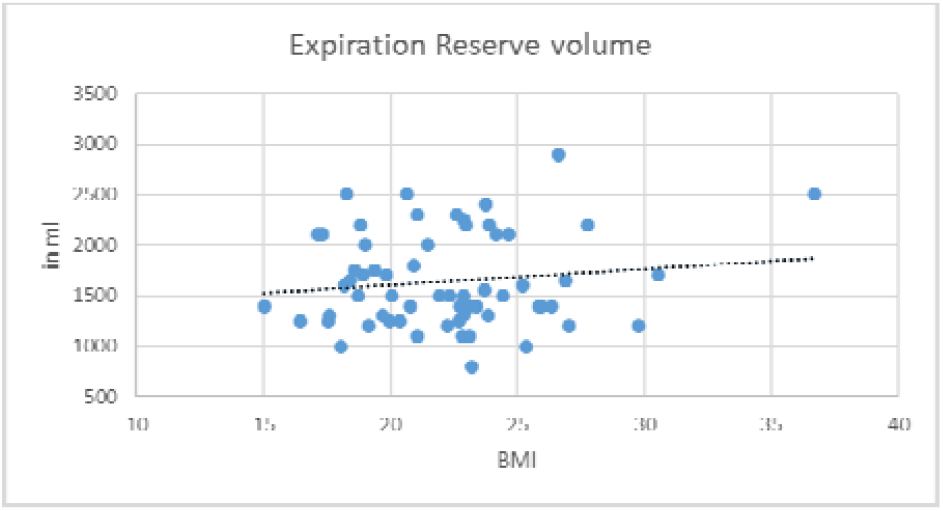

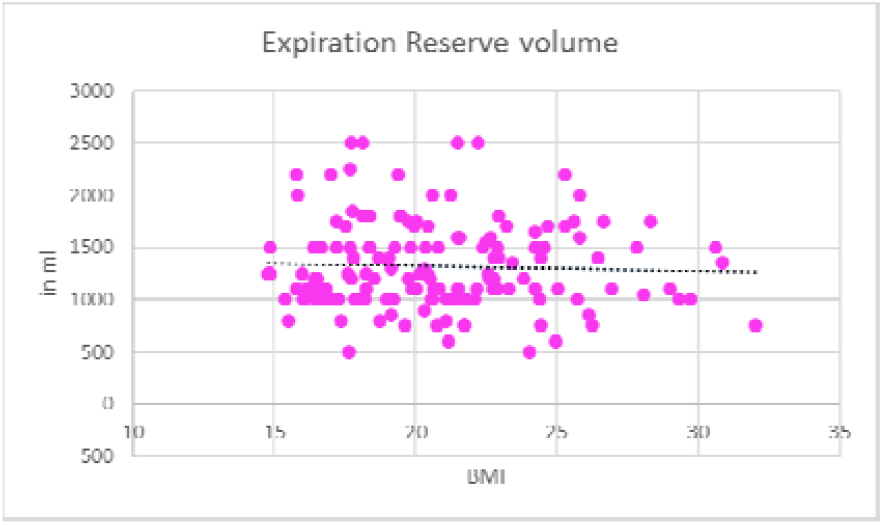
  d. Vital Capacity Standing (Table 10, Figure 14, Figure 15)

| R- value ( vs BMI) | Males | Females |
| --- | --- | --- |
| VC Standing | 0.0536 | 0.0412 |

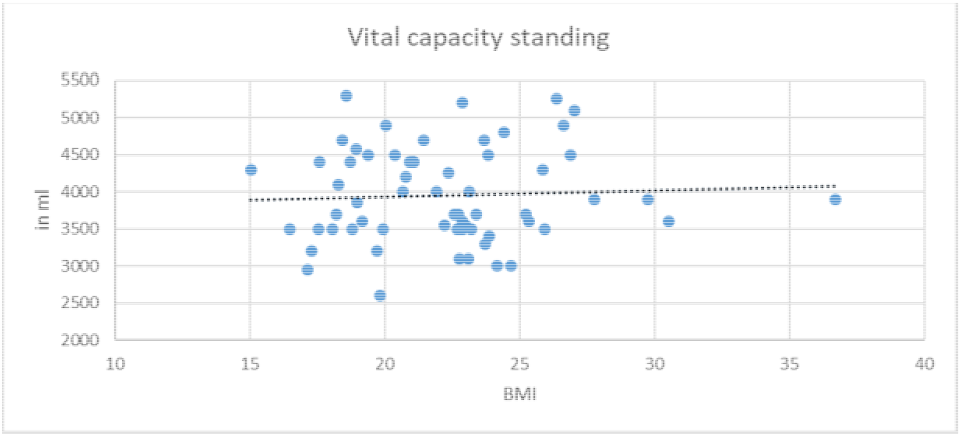

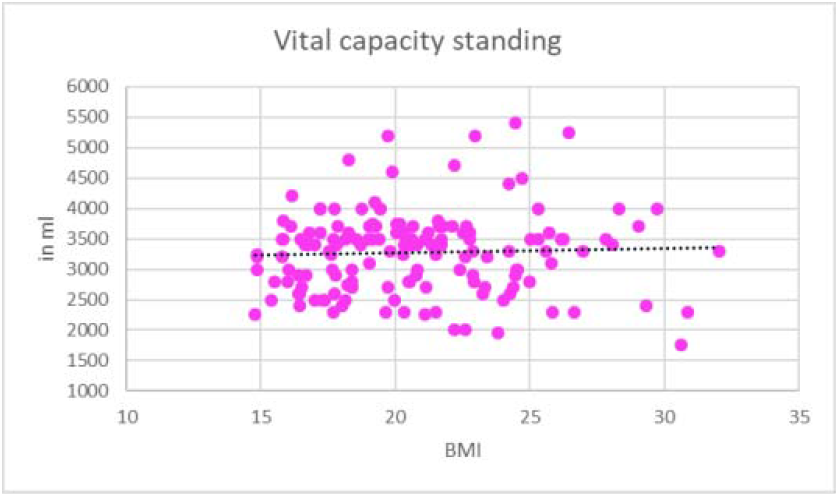
  e. Vital Capacity Sitting (Table 11, Figure 16, Figure 17)

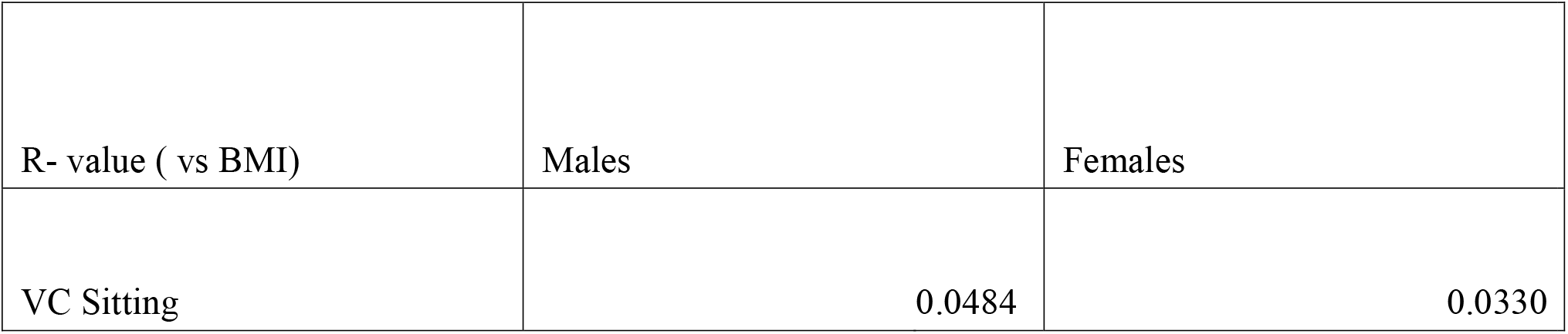

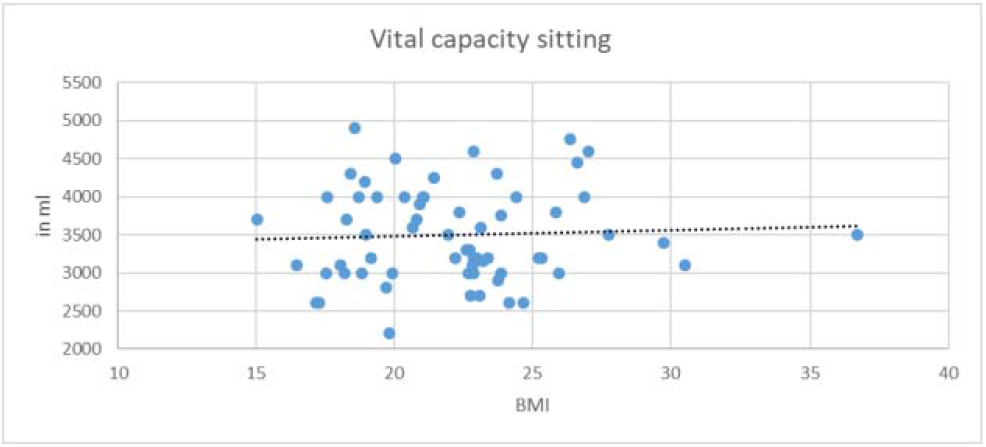

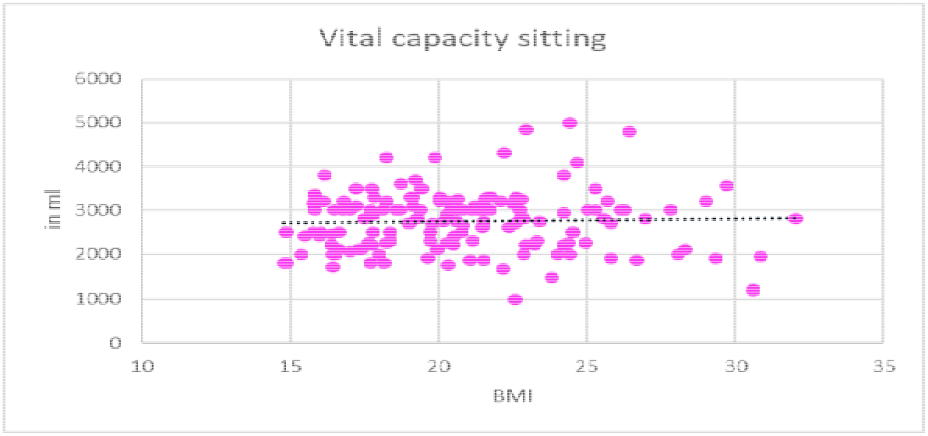
  f. Maximum Expiratory Pressure (Table 12, Figure 18, Figure 19)

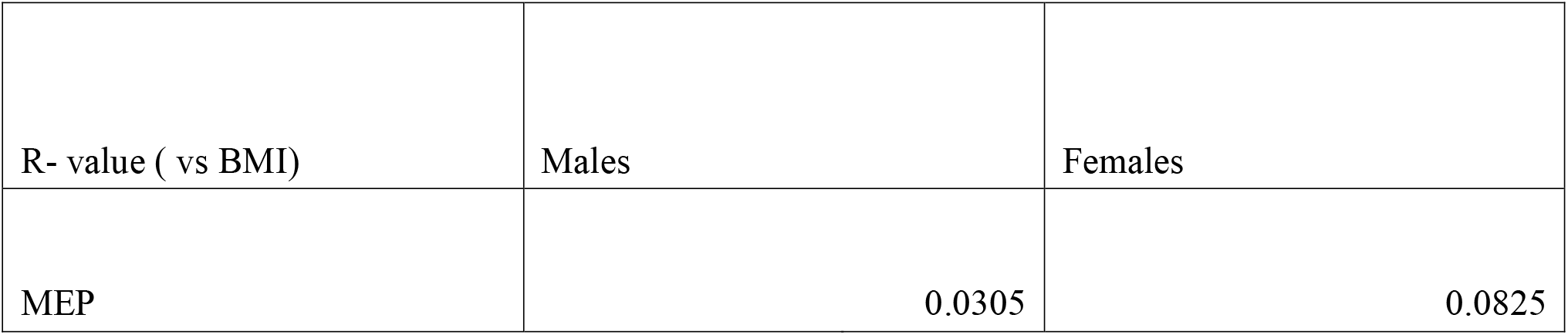

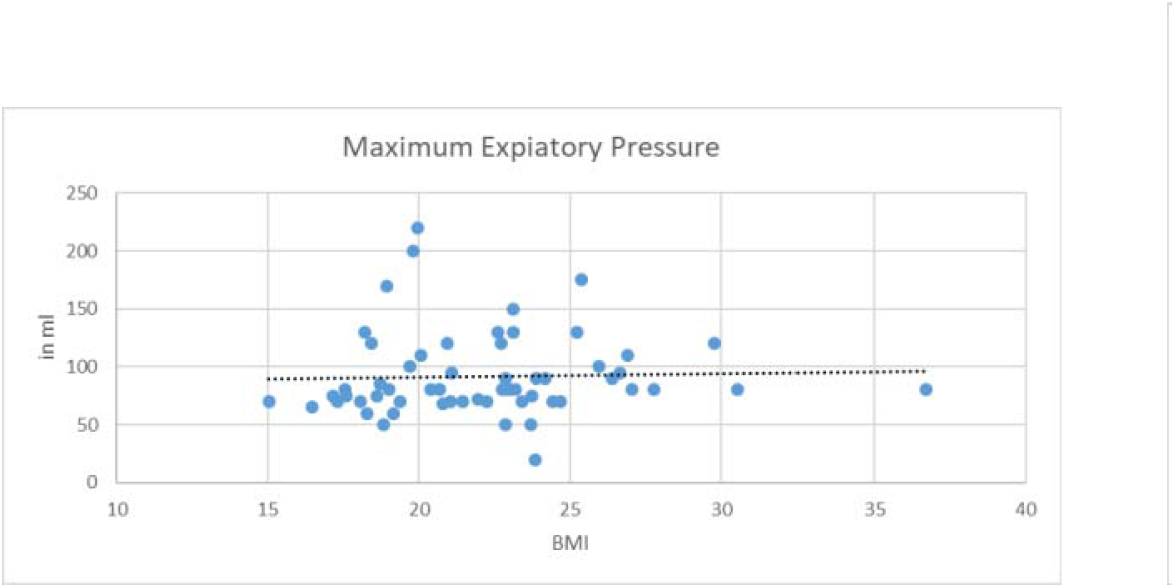

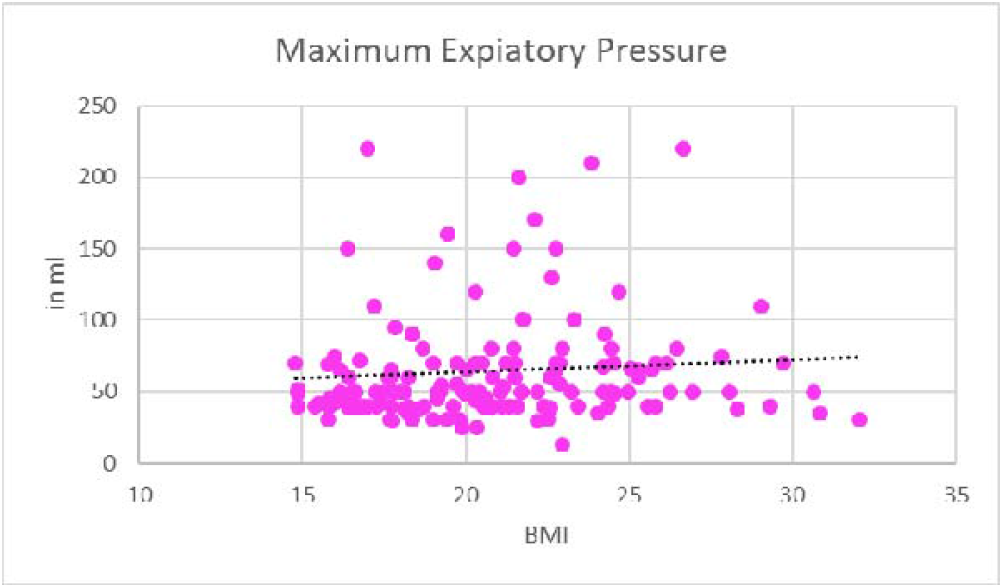

## Discussion

It is evident from Table 1 and figure 1 that values of Tidal volume is in the range of tidal volumes noted in standard physiology textbooks. Males have less Tidal volume than females and the difference is statistically significant. Comparison of Pearson Correlations of BMI and all respiratory parameters between males and females for Tidal volume (Table 7, Figure 8, Figure 9) show very weak negative correlation between BMI and TV In both males and females, which is not significant and is weaker in males than in females to provide sufficient evidence that BMI cannot be correlated with TV and has no effect on it whatsoever.

By Table 2 and figure 2 note can be made that values of Inspiratory Reserve volume are similar as noted in standard physiology textbooks. Also, It can be clearly noticed that males have statistically significant higher IRV than females. Which sums up as males had higher Inspiratory Capacity (TV was slight less but they make up for it in IRV) than females. Comparison of Pearson Correlations of BMI and all respiratory parameters between males and females for Inspiratory reserve volume (Table 8, Figure 10, Figure 11) show very weak positive correlation between BMI and IRV in both males and females which is not significant and is weaker in females than in males, w This provides evidence that BMI cannot be correlated with IRV and has no effect on it whatsoever

Table 3 and figure 3 demonstrate that Expiratory Reserve volume we found is similar as in standard physiology textbooks. The graphs provide sufficient evidence that Males have a statistically significant higher ERV than females, which sums up as higher Expiratory Capacity (slight lesser TV but they make up for it in ERV) than females. Comparison of Pearson Correlations of BMI and all respiratory parameters between males and females for Expiratory Reserve Volume (Table 9, Figure 12, Figure 13), a weak negative correlation was found between BMI and ERV in females and weak positive correlation in males. Whatsoever, the correlation is not strong enough to be deemed significant. which proves that ERV and BMI cannot be correlated and that the later has no impact on the formal.

Table 4 and Figure 4 demonstrate that Vital capacity while standing is in the normal range denoted in standard physiological textbooks. Males have statistically significant higher VC while standing then females, and also, it can also be noticed here that high IRV and ERV in males make up for the lesser TV to give higher VC. Comparison of Pearson Correlations of BMI and all respiratory parameters between males and females for Vital Capacity Standing (Table 10, Figure 14, Figure 15) show, very weak positive correlation between BMI and VC standing which is not significant. It was weaker in females than in males and it provides enough evidence that BMI cannot be correlated with VC standing and has no effect on it whatsoever.

With the help of Table 5 and Figure 5, we can see that Vital capacity while sitting is in the normal range denoted in standard physiology textbooks and it also shows that males have statistically significant higher VC while sitting then females. As seen in VC while standing, sitting position also shows the same trend where higher IRV and ERV makes up for the lesser TV. Comparison of Pearson Correlations of BMI and all respiratory parameters between males and females for Vital Capacity Sitting (Table 11, Figure 16, Figure 17) show, very weak positive correlation between BMI and VC sitting. This amount of correlation is not significant. It was weaker in females than in males and provides sufficient evidence that BMI cannot be correlated with VC sitting and has no effect on it whatsoever

Table 6 and figure 6 denotes the Maximum expiratory pressure which was found to be in normal range denoted in standard physiological textbooks. It is also visible here that males have statistically significant higher MEP then females which can be attributed to more muscular framework and male anatomy in general. Comparison of Pearson Correlations of BMI and all respiratory parameters between males and females for Maximum Expiratory Pressure (Table 12, Figure 18, Figure 19) show, very weak positive correlation between BMI and MEP. Weaker in males than in females this degree of correlation is not significant. It provides sufficient evidence that BMI cannot be correlated with MEP and has no effect on it whatsoever.

A research conducted to find Linear relationship between BMI and Vital Capacity and Total Lung Capacity found that, although a significant correlation can be found between both, lung parameters remain normal ranges even for morbidly obese patients which can be noticed in our study. (13) While Another study done on a generally age distributed small sample size of females showed a positive correlation between BMI and IRV similar to our study but found negative correlation between ERV and BMI. (14) A Novel research using predicting equations shown that if participants in a sample are grouped with taking their degree of obesity as distributing factor, Prediction of values can be done with high accuracy and high precision with high level of confidence and less error rate. This also proves that BMI and Lung capacities can be correlated. (15) A study found that FRC and ERV are negatively correlated with BMI and decrease with increasing obesity leading to respiration towards their residual volume (RV). It also showed that obesity has modest effect on TLC and RV. The results are similar to our study as we also found negative correlation between ERV and BMI in females, and although it was positive in males, it was very small and weak. (16) One of the research article found out same as other articles including ours that ERV and FRC decreases with increasing BMI. They also showed that IC increases with increasing BMI and the results align with our study as we also found positive correlation both in males and females between IRV and BMI. But they found an inverse negative correlation between BMI and VC whereas we found a very weak correlation. (17) A cohort study on 13 children weighting 1.47-3.00 times the ideal weight for age and height, showed that when comparison is carried out with normal weighted children, all of the following namely, Forced expiratory volume and forced expiratory flow rate and also, MVV (Maximum Voluntary Ventilation) decrease except Residual Volume, RV/TLC (Total Lung Capacity), VE (Minute Ventilation), REE (Resting energy expenditure) increases. All other Lung volumes and capacities remain the same, which aligns with the result we found as the level of Pearson correlation that we found is too weak. (18) A study found dramatically decreased in ERV and Functional Residual Volume (FRC) when BMI and obesity was increasing. It is similar to our results although we found not a very gross change as depicted in the study. (19)

## Conclusion

With the help of the results found and the comparison made with other studies, it can be found that BMI does not have a significant impact on respiratory parameters and maintaining a healthy body weight and BMI do certainly helps in respiration and oxygenation which at the end of the day only increases one’s capacity to tolerate oxygen stress and work better and more efficiently but is not the only factor affecting it. We also found that there is not much difference between normal values of lung volumes and capacities between European people and South Asian (Indian) population and he figures that are given in standardized books are as relevant in India as they are in other countries.

## Data Availability

All data produced in the present study are available upon reasonable request to the authors

## Acknowledgement

We, the authors would like acknowledge the following Medical Students of Smt. NHLMMC for their efforts and work in maintaining decorum and oversee the data collection process along with authors and Physiology department to see the correct techniques were utilized while doing the experiment and that correct values were recorded.

- Dr. Kshitish Brahmachari
- Dr. Vidit Jain
- Dr. Devasya Patel
- Dr. Mohit Patel
- Dr. Bhairavi Halani
- Dr. Falak Saiyed
- Dr. Khushi Dave

We would also like to thank the Physiology Department of SMT. NHLMMC to assist us and help us through research project and guide us towards a better understanding of spirometry and Human Physiology.

